# Imprecision in tuberculosis infection outcomes; implications for non-inferiority vaccine trials

**DOI:** 10.1101/2025.06.19.25329919

**Authors:** Daniel J Grint, Richard G White, Gavin Churchyard, Andrew Fiore-Gartland, Molebogeng X Rangaka, Alberto L. Garcia-Basteiro, Frank Cobelens

## Abstract

**Introduction:** Randomised trials comparing new vaccines against tuberculosis for use in neonates and infants, for whom Bacille Calmette-Guérin (BCG) vaccination is established practice, are using tuberculosis infection as the primary endpoint in a non-inferiority design. Markers of tuberculosis infection have imperfect sensitivity and specificity. Flaws in the non-inferiority trial design typically bias towards the null, which may result in falsely declaring non-inferiority.

**Methods:** We conducted a statistical simulation study to assess the impact of imperfect markers of tuberculosis infection on the interpretation of tuberculosis vaccine trials testing a non-inferiority hypothesis of an infection primary outcome in a two-arm randomized comparison. Data were generated in three 2-year cumulative risk of tuberculosis infection scenarios (2%, 5%, and 8%). The specificity of tests of tuberculosis infection was assumed to range from 100% to 85%, while the sensitivity was assumed to range from 100% to 64%. Log-binomial regression was used to estimate the relative risk of tuberculosis infection.

**Results:** With 100% sensitivity and specificity, type-I and type-II error were both approximately equal to the expected values (2.5% and 80%, respectively) in all three cumulative tuberculosis risk scenarios. With modest deviations from perfect sensitivity and specificity (95% for both), the risk of falsely declaring non-inferiority was 96.8%, 53.2%, and 27.8% in the 2%, 5%, and 8% cumulative tuberculosis risk infection scenarios, respectively.

**Discussion:** Tuberculosis vaccine non-inferiority trials using an infection primary outcome must be designed and interpreted accounting for the specificity of the tools used to measure infection, otherwise they risk declaring non-inferiority by default.

**Key messages:**

- We conducted a statistical simulation study to assess the impact of imperfect sensitivity and specificity, in the primary outcome definition of tuberculosis infection, in vaccine trials testing a non-inferiority hypothesis.
- With only modest departures from perfect specificity in tuberculosis infection markers, the risk of falsely declaring non-inferiority is substantial.
- Vaccine trials testing a non-inferiority hypothesis with an infection primary outcome must account for the imprecision in the tools used to define the outcome, otherwise vaccines may be falsely declared non-inferior.

## Introduction

Non-inferiority trial designs are commonly used in the assessment of new treatments and other interventions where there is an established therapeutic standard of care, and are also becoming increasingly common for trials of new vaccines.^1 2 3^

Tuberculosis (TB) is the most common cause of death due to a single infectious agent globally.^4^ The only available TB vaccine, Bacille Calmette-Guérin (BCG), has existed for over 100 years and provides 80% (95% confidence interval, 34-94%) protection against severe forms of TB disease in children but inconsistent protection against pulmonary TB at later age.^5^ The World Health Organisation (WHO) has identified new vaccines against TB as a global health priority, and several candidates are in clinical development.^6^

Presently, there are several ongoing randomised trials comparing new vaccines against TB for use in neonates and infants, for whom BCG vaccination is established practice.^6^ The WHO’s Product Profile Characteristics (PPC) for a new TB vaccine for use in this age group stipulates that it should have superior efficacy for prevention of TB disease as compared to BCG, improved safety in neonates and infants with innate or acquired immunodeficiency, including HIV infected infants, or improved manufacturing scalability and lower costs.^7^

In addition, TB disease has a low background incidence and long incubation periods, making trials with a prevention of disease (PoD) outcome large, long and costly. As TB infection occurs much more commonly than disease, trials have sought to adopt prevention of TB infection (PoI) as a surrogate primary outcome endpoint.^8 9^ Infection is generally measured as interferon-gamma release assays (IGRA) conversion that measures an adaptive immune response to *M. tuberculosis*-specific antigens. Although there is no gold standard for establishing TB infection, false-negative IGRA results can occur in people with bacteriologically confirmed TB.^10^ False-positive IGRA results may occur insofar that TB infection defined by lower IGRA cut-off values are known to progress to TB disease less often than TB infection based on higher IGRA cut-off values.^11^

Some current trials are using a non-inferiority trial design in comparing the efficacy of new vaccines to that of BCG with TB infection as the primary endpoint. Non-inferiority trials pose a risk of falsely declaring non-inferiority under certain conditions relating to trial conduct and the choice of primary outcome.^12^ This is because in a non-inferiority trial, flaws in trial design typically bias towards the null, in favour of non-inferiority. This is a well-established principle for drug trials, where regulators provide extensive guidance on endpoint selection and trial management.^13^ The same principles have not yet been clearly established for vaccine trials, which differ from drug trials in that the primary outcome is usually specified as a relative risk rather than a risk difference, which is interpreted as protective efficacy.^14^ In vaccine studies a relative risk is preferred because, contrary to a risk difference, it provides a measure of effect that is independent of the underlying disease incidence.

Since IGRA have suboptimal diagnostic accuracy for TB infection,^10 11^ TB infection outcomes reported by trials may be dominated by false test positives rather than true TB infections, particularly in low infection incidence populations. As vaccine trials are typically analysed using relative effect measures, false test positive outcomes may diminish the true relative difference between control and experimental vaccines and result in falsely declaring non-inferiority (Figure 1). Conversely, false test negative outcomes may diminish the statistical type-II error for relative effect measures in a non-inferiority vaccine trial.^15^

**Figure 1.**
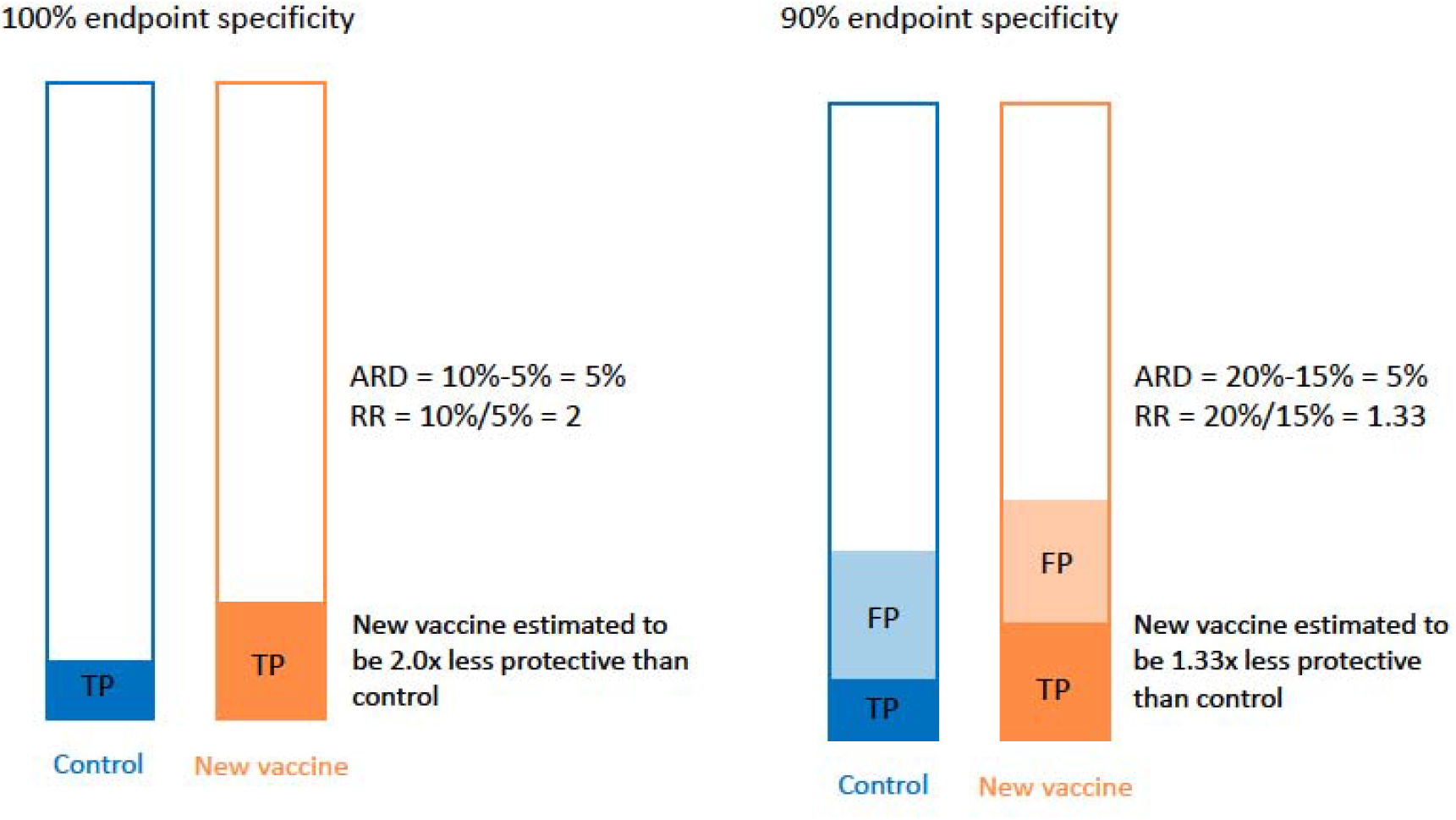
Hypothetical example showing the effect of incomplete specificity of the measurement of the endpoint in a vaccine trial on the estimated absolute risk difference (ARD) and estimated relative risk (RR). Bars represent the full study population for each trial arm, shaded areas the proportion positive on a test. TP: true positives. FP: false positives. The relative risk is biased towards 1, whereas the absolute risk difference is not affected. The vaccine’s efficacy is commonly interpreted as 1 - RR

This simulation study assessed the impact of imperfect sensitivity and specificity of TB infection diagnostics on type-I and type-II error in simulated non-inferiority trials.

## Methods

We conducted a statistical simulation study to assess the impact of imperfect sensitivity and specificity in the outcome definition of TB infection in vaccine trials testing a non-inferiority hypothesis. We aimed to demonstrate the impact of imperfect outcome sensitivity and specificity on the type-I and type-II error of non-inferiority vaccine trials.

Data were generated from the binomial distribution with probability equal to the assumed risk of TB infection following vaccination. Data were generated from hypothetical non-inferiority trials under three scenarios for the risk of TB infection in the control arm following vaccination, reflecting three different levels of cumulative incidence of IGRA conversion during two years of follow-up (2%, e.g. rural East Africa; 5% e.g. urban East Africa, 8%, South Africa).^16^

The simulated sample size for each scenario was informed by standard statistical formulae based the normal distribution approximation,^17 18^ and refined in simulations to have approximately 80% type-II error to test the null hypothesis that the upper confidence interval for the relative risk is not greater than 1.25, assuming a one-sided type-I error of 2.5% (Table 1). We selected this non-inferiority margin assuming that a vaccine 1.25 times less efficacious than BCG in protecting neonates and infants against TB infection would not be considered non-inferior regardless of possible advantages with regard to safety or cost.

**Table 1.** Parameters for data generation.

| Tuberculosis infection risk | Total sample size | Non-inferiority margin | Type-II error analysis |  | Type-I error analysis |  |
| --- | --- | --- | --- | --- | --- | --- |
|  |  |  | Relative risk | Expected type-II error | Relative risk | Expected type-I error (one-sided) |
| 0.02 | 32,000 | 1.25 | 1 | 80% | 1.25 | 2.5% |
| 0.05 | 12,000 | 1.25 | 1 | 80% | 1.25 | 2.5% |
| 0.08 | 7,200 | 1.25 | 1 | 80% | 1.25 | 2.5% |

The trial populations were generated with 2,000 replications to give an approximate Monte Carlo standard error of <1% for the estimated type-I and type-II error.^19^

Following data generation of the trial populations and *true* TB infection cases, *observed* TB infections were computed from a binomial distribution applying varying degrees of test sensitivity and specificity.

The targets of the analysis were the type-I and type-II error of the test of the null hypothesis of non-inferiority of the experimental vaccine compared to the control. For both targets, the method of analysis was a log-binomial regression model computing the relative risk of TB infection.

For analysis of type-I error, data were generated under a *true* relative risk of 1.25. Type-I error was computed as the proportion of simulated trials where the upper limit of the confidence interval for the relative risk comparing the experimental vaccine to the control vaccine was <1.25. In other words, type-I error was calculated as the proportion of simulated trials where non-inferiority was demonstrated, when the true risk of TB infection was 1.25 times higher with the experimental vaccine.

For analysis of type-II error, data were generated under a *true* relative risk of 1. Type-II error was computed as the proportion of simulated trials where the upper limit of the confidence interval for the relative risk comparing the experimental vaccine to the control vaccine was ≤1.25. In other words, type-II error was calculated as the proportion of simulated trials where non-inferiority was demonstrated, when there was truly no difference in the efficacy of the experimental and control vaccines.

Analyses were conducted under the following assumptions on the diagnostic accuracy of the TB infection: sensitivity and specificity both 100%, sensitivity 100% and specificity 95%, sensitivity 95% and specificity 100%, sensitivity and specificity both 95%, and sensitivity 95% and specificity 98% (Table 2).^11 20^ As an extreme case we also included sensitivity 64% and specificity 85%. These values were calculated from a meta-analysis of cohort studies from low-incidence countries that included immigrants from high-incidence countries.^10^

**Table 2.** Sensitivity and specificity combinations of TB infection markers used in data generation.

| Sensitivity | Specificity |
| --- | --- |
| 100% | 100% |
| 100% | 95% |
| 95% | 100% |
| 95% | 95% |
| 95% | 98% |
| 64% | 85% |

Data generation and statistical analysis was performed using R version 4.3.3 and R Studio 2023.12.0 build 369. The code used to generate the data and perform all analyses is available at https://github.com/dgrint/TB-Vacc.

## Results

### Analysis of type-I error

Type-I error here refers to the proportion of simulated trials where non-inferiority was demonstrated, when the true risk of TB infection was 25% higher with the experimental vaccine than the control.

Results of the simulated type-I error analysis are summarised in Figure 2 and Table 3. When sensitivity and specificity of the diagnostic tools are both 100%, type-I error is approximately equal to 2.5% for all three true TB infection risk scenarios.

**Figure 2.**
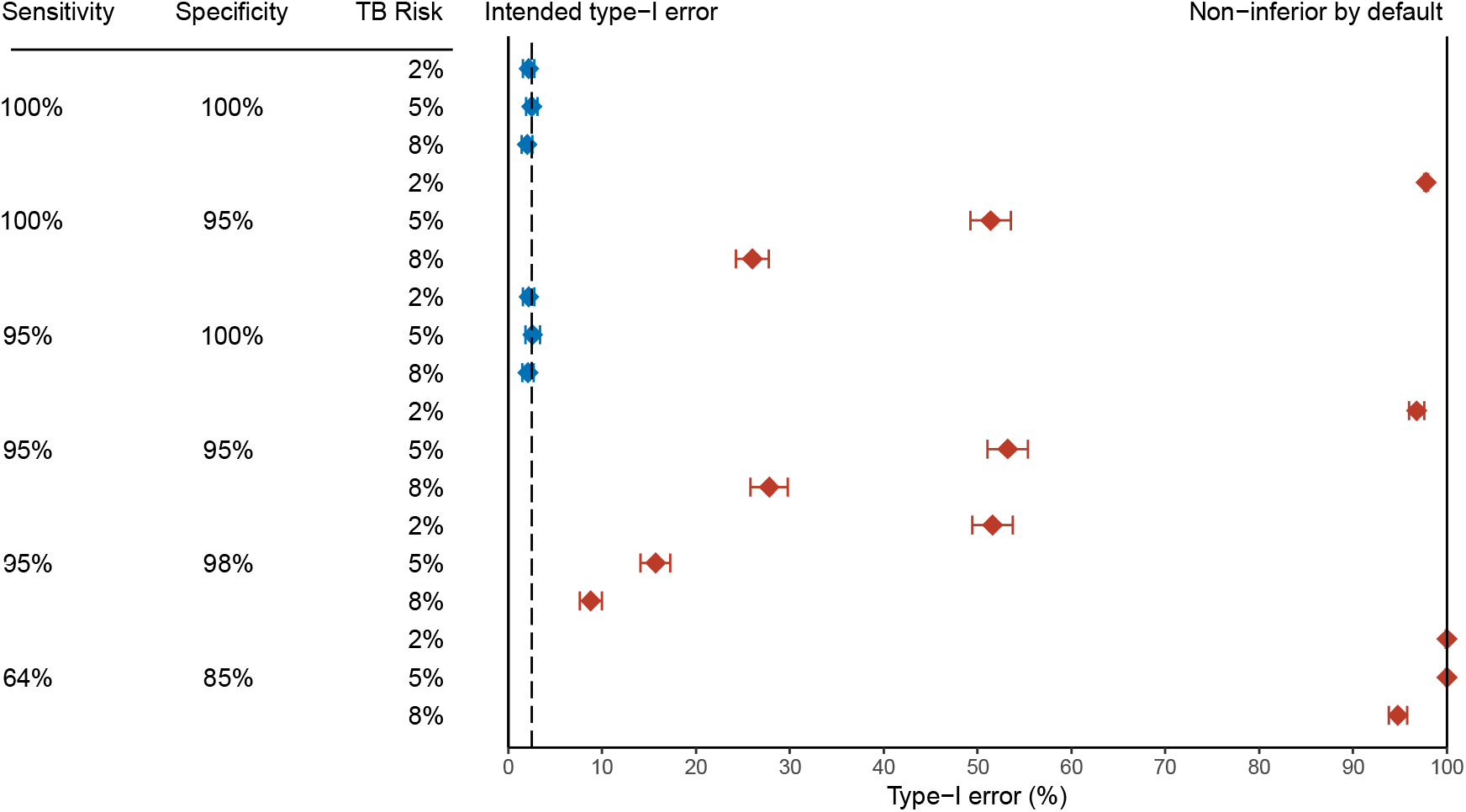
Probability of falsely declaring non-inferiority (Type-I error) for different combinations of sensitivity and specificity of the measurement of the endpoint, and the cumulative infection incidence, in 2,000 simulated tuberculosis vaccine trials with an infection endpoint for which true risk is 25% higher with the experimental vaccine than with the control. Horizontal bars: 95% Monte Carlo error bounds. Blue diamonds: combinations for which the observed Type-I error is consistent with 2.5% (intended Type-I error). Red diamonds: combinations for which the observed Type-I error is larger than 2.5%. At 100% (right margin) non-inferiority will be declared by default (in all trials).

**Table 3.**
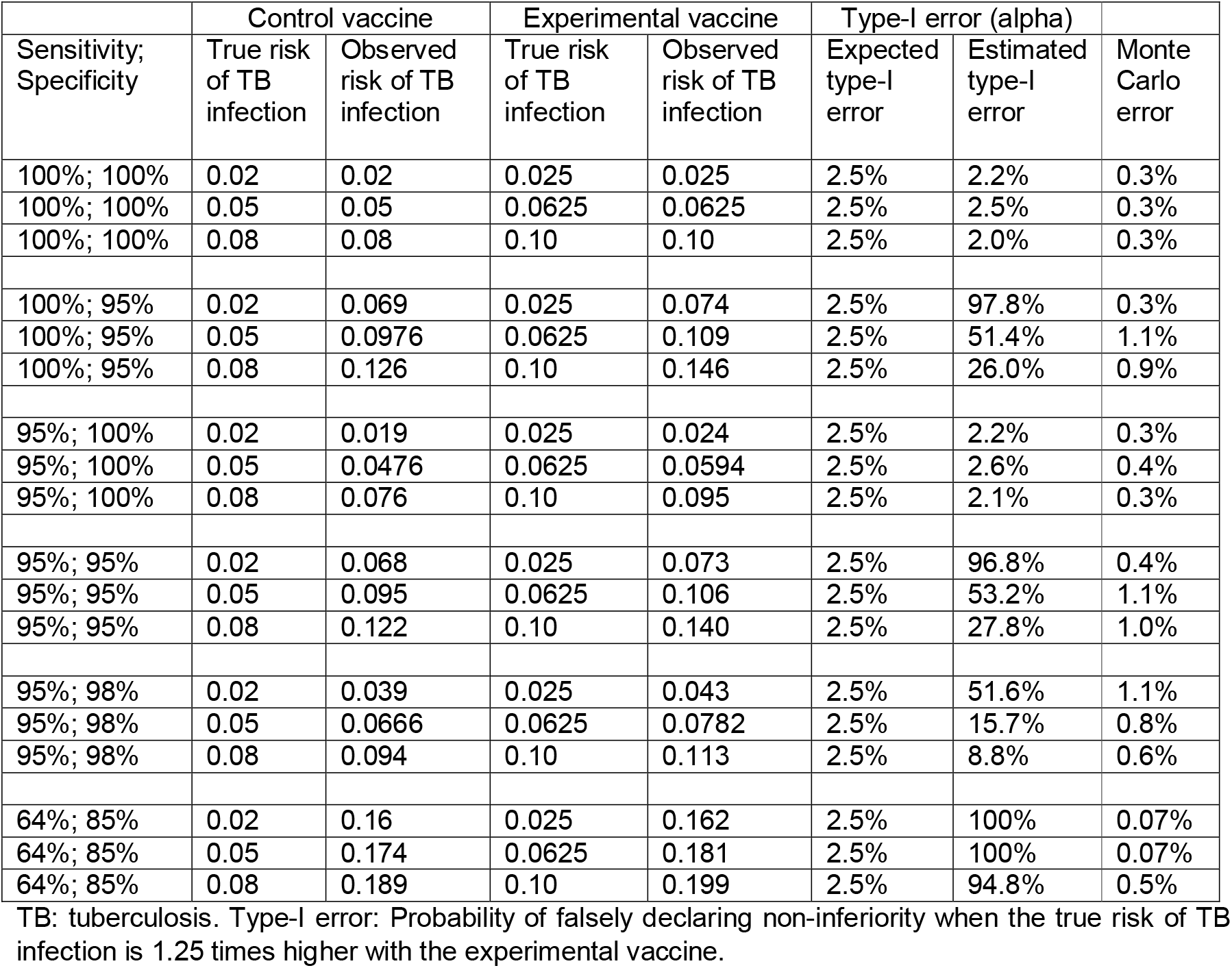
Impact of imperfect sensitivity and specificity of TB infection outcome on type-I error.

With 100% sensitivity and 95% specificity, type-I error increases to 97.8% when the true TB infection risk is 2%. Meaning non-inferiority would be falsely declared in 97.8% of trials where the true risk of TB infection was 1.25 time higher with the experimental vaccine. Type-I error is 51.4% and 26.0% for true TB infection risks of 5% and 8%, respectively, with similar results when sensitivity and specificity are both 95% (96.8%, 53.2%, and 27.8%, respectively).

Sensitivity of 95% combined with perfect specificity had only a modest impact on type-I error. However, even a modest reduction in specificity to 98% resulted in inflated type-I error for all three scenarios (51.6%, 15.7%, and 8.8%, respectively).

In the extreme case of 64% sensitivity and 85% specificity, type-I error was inflated to 100% for the 2% and 5% TB infection risk scenarios and 94.8% for the 8% scenario. Meaning virtually all trials would demonstrate non-inferiority in this scenario.

### Analysis of type-II error

Type-II error here refers to the proportion of simulated trials where non-inferiority was demonstrated, when the true risk of TB infection was the same with experimental vaccine and the control. Greatly inflated type-II error indicates an overwhelming likelihood of demonstrating non-inferiority.

Results of the simulated type-II error analysis are summarised in Figure 3 and Table 4. When sensitivity and specificity of the diagnostic tools are both 100% the observed risk of TB infection matches the true risk of TB infection, consequently type-II error is approximately equal to 80% for all three true TB infection risk scenarios.

**Figure 3.**
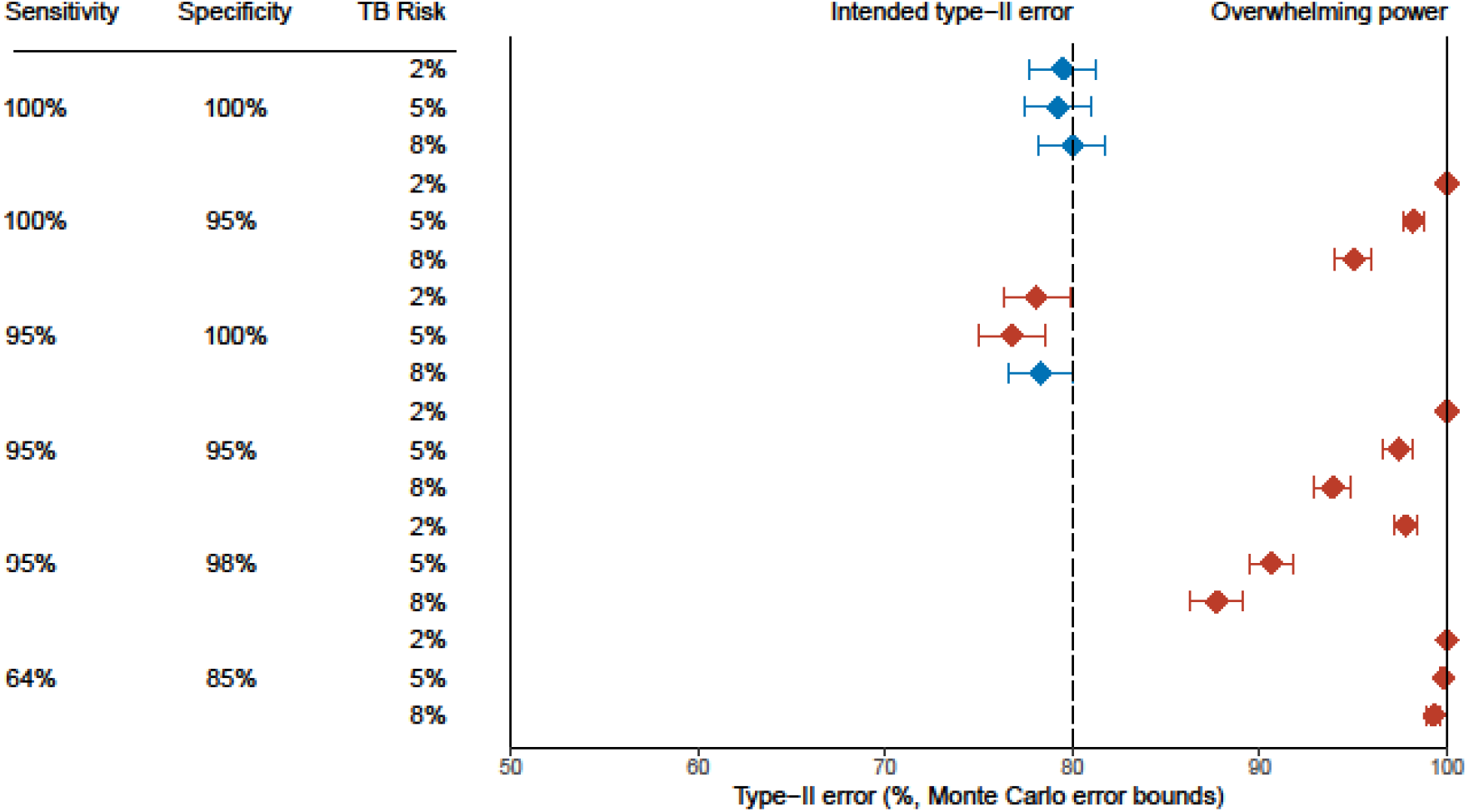
Probability of correctly declaring non-inferiority (Type-II error) for different combinations of sensitivity and specificity of the measurement of the endpoint, and the cumulative infection incidence, in 2,000 simulated tuberculosis vaccine trials with an infection endpoint for which true risk is 25% higher with the experimental vaccine than with the control. Horizontal bars: 95% Monte Carlo error bounds. Blue diamonds: combinations for which the observed Type-II error is consistent with 80% (intended Type-II error). Red diamonds: combinations for which the observed Type-II error deviates from 80%. At 100% (right margin) non-inferiority will be declared by default.

**Table 4.** Impact of imperfect sensitivity and specificity of TB infection outcome on type-II error.

|  | Control vaccine |  | Experimental vaccine |  | Type-II error (beta) |  |  |
| --- | --- | --- | --- | --- | --- | --- | --- |
| Sensitivity;<br>Specificity | True risk<br>of TB | Observed<br>risk of TB | True risk<br>of TB | Observed<br>risk of TB | Expected<br>type-II | Estimated<br>type-II | Monte<br>Carlo |

|  | infection | infection | infection | infection | error | error | error |
| --- | --- | --- | --- | --- | --- | --- | --- |
| 100%; 100% | 0.02 | 0.02 | 0.02 | 0.02 | 80% | 79.5% | 0.9% |
| 100%; 100% | 0.05 | 0.05 | 0.05 | 0.05 | 80% | 79.2% | 0.9% |
| 100%; 100% | 0.08 | 0.08 | 0.08 | 0.08 | 80% | 80.0% | 0.9% |
| 100%; 95% | 0.02 | 0.069 | 0.02 | 0.069 | 80% | 100% | 0.07% |
| 100%; 95% | 0.05 | 0.0976 | 0.05 | 0.0975 | 80% | 98.2% | 0.3% |
| 100%; 95% | 0.08 | 0.126 | 0.08 | 0.126 | 80% | 95.0% | 0.5% |
| 95%; 100% | 0.02 | 0.019 | 0.02 | 0.019 | 80% | 78.1% | 0.9% |
| 95%; 100% | 0.05 | 0.0476 | 0.05 | 0.0475 | 80% | 76.8% | 0.9% |
| 95%; 100% | 0.08 | 0.076 | 0.08 | 0.076 | 80% | 78.3% | 0.9% |
| 95%; 95% | 0.02 | 0.068 | 0.02 | 0.068 | 80% | 100% | 0.07% |
| 95%; 95% | 0.05 | 0.095 | 0.05 | 0.095 | 80% | 97.4% | 0.4% |
| 95%; 95% | 0.08 | 0.122 | 0.08 | 0.122 | 80% | 93.9% | 0.5% |
| 95%; 98% | 0.02 | 0.039 | 0.02 | 0.039 | 80% | 97.8% | 0.3% |
| 95%; 98% | 0.05 | 0.0665 | 0.05 | 0.0665 | 80% | 90.6% | 0.6% |
| 95%; 98% | 0.08 | 0.0945 | 0.08 | 0.0942 | 80% | 87.7% | 0.7% |
| 64%; 85% | 0.02 | 0.16 | 0.02 | 0.16 | 80% | 100% | 0.07% |
| 64%; 85% | 0.05 | 0.174 | 0.05 | 0.175 | 80% | 99.8% | 0.1% |
| 64%; 85% | 0.08 | 0.189 | 0.08 | 0.189 | 80% | 99.3% | 0.2% |

TB: tuberculosis. Type-II error: Probability of declaring non-inferiority when the true risk of TB infection is the same with the control and experimental vaccine.

When specificity is below 100% the observed risk of TB infection is considerably higher than the true risk of TB infection. When 1-specificity is greater than the true risk of TB infection, there are more false positives than true positives in the observed risk of TB infection.

With 100% sensitivity and 95% specificity, type-II error is ≥95% in all scenarios. With 95% sensitivity and 95% specificity, type-II error remains ≥93% for all scenarios, indicating that imperfect sensitivity does not counterbalance imperfect specificity.

Sensitivity of 95% combined with perfect specificity has only a modest impact on type-II error, with type-II error reduced just below 80% in all three scenarios. However, even a modest reduction in specificity to 98% results in inflated type-II error for all three scenarios, and considerably so when the true risk of TB infection is low.

In the extreme case of 64% sensitivity and 85% specificity, type-II error is inflated to approximately 100% for all three TB infection risk scenarios.

## Discussion

This study demonstrated that even modest departures from perfect specificity in tests of TB infection outcomes result in vastly inflated type-I and type-II error in trials using TB infection as the primary outcome. Consequently, the risk of declaring non-inferiority by default is high, particularly when the true risk of TB infection over the follow-up period, in this case 2-years, is similar to or less than the 1-specificity of the diagnostic test.

This study assessed the impact of modest, as well as extreme, departures from perfect diagnostic performance. There is no established gold standard diagnostic to define TB infection. The sensitivity of IGRA has generally been assessed among patients with bacteriologically confirmed TB and estimated at close to 95% for the latest generation assays.^20^ Various approaches have been used to assess the specificity of IGRA for TB infection for use in vaccine trials. The estimate that matters most is that for predicting TB disease occurring over a two-year period.^21^

A cohort study that followed South African infants in a high TB incidence setting 6-24 months post QuantiFERON testing showed close to 95% specificity for TB disease at the manufacturer recommended cut-off of 0.35 IU/ml and close to 98% specificity at cut-off 4.0 IU/ml.^11^ However, the specificity calculated for any IGRA test at manufacturer recommended cut-off from a meta-analysis of cohort studies from (mainly) low-incidence countries that included immigrants from high-incidence countries of all ages was only 85%.^10^ The difference may be due to differences in age groups, which would make the higher specificity estimate more applicable for our study. It may also reflect technical variations in performing the test in field conditions, and differences in incidence of TB infection. In multi-country TB vaccine trials IGRA may therefore have a specificity closer to our extreme case of 85%. The sensitivity calculated from the meta-analysis of cohort studies was 64%.^10^ Although this low estimate may partially reflect bias due to infections occurring between IGRA testing and diagnosis of TB disease,^21^ we included it in our simulations as an extreme case.

The probability of falsely declaring non-inferiority was dependent on the cumulative risk of TB infection in the population where the trial was conducted. Most TB vaccine trials will be conducted in populations with 5 to 8% cumulative infection risk over two years trial follow-up. Our simulations suggest that the probability of falsely declaring non-inferiority was lowest for the highest incidence setting, but still inflated tenfold (26%) when specificity was 95%. In the extreme case of 85% specificity, the probability of falsely declaring non-inferiority was 94.8% even at the highest background incidence of TB infection.

We assumed that TB infection status was measured only once, at the end of trial follow-up. In reality, trials may do repeated IGRA testing, e.g. at six-month intervals. In such cases, false-positive IGRA results will occur at every test round and the overall specificity may be even lower, further increasing the risk of falsely declaring non-inferiority. On the other hand, if sustained infection, requiring two or more serial positive tests, is a requirement to determine infection, the specificity of the endpoint may increase.^9^

It could be argued that the results presented here are not surprising as the observed TB infection risk is different from the true TB infection risk that was used to inform the sample size calculations. The analysis performed on the observed risk of TB infection is then a departure from the study design, leading to the inflated type-I and type-II error rates shown here. Indeed, researchers must account for the precision of the tools measuring their primary outcomes when designing their trials. However, when the proportion false-positive results due to the diagnostic tool is close to or lower than the risk of the disease itself, the validity of the tool as a measure of defining the primary outcome must be called into question.

The considerations underlying our analyses may also have implications for non-inferiority trials with TB disease as the endpoint. Trials among neonates and infants in whom bacteriological confirmation of the diagnosis can be challenging often have composite clinical endpoints, therefore diagnostic specificity of the outcome in these trials may be compromised. The extent to which non-inferiority may be falsely declared then also depends on the proportion of the trial participants who are assessed for TB disease, which is usually based on symptom screening. For a given diagnostic specificity, the number of false-positive diagnosis will increase with the proportion trial participants who are tested for the disease endpoint, as will the probability of falsely declaring non-inferiority.

Vaccine non-inferiority trials using a marker of infection as the primary outcome must be designed and interpreted accounting for the precision of the tools used to measure infection, otherwise they risk declaring non-inferiority by default. Until there is a test for TB infection with higher specificity, the use of the non-inferiority trial design for TB vaccine prevention of infection outcomes is not recommended.

## Data Availability

The simulated data for this study are available through the github link given in the methods section.

https://github.com/dgrint/TB-Vacc

## Declarations

### Ethics approval

Ethics approval is not required for statistical simulation studies.

### Author contributions

FC and DJG designed the study. DJG wrote the code and performed all statistical analysis. DJG and FC drafted the manuscript. All other authors contributed substantially to revising and refining the manuscript.

### AI tools

No AI tools were used in preparing this manuscript.

